# The effect of level of injury on diabetes incidence and mortality after spinal cord injury – a longitudinal cohort study

**DOI:** 10.1101/2023.05.23.23290398

**Authors:** Sven Hoekstra, Michelle Trbovich, Wouter Koek, Michael Mader, Marzieh Salehi

## Abstract

**Objective:** Persons with spinal cord injury (SCI) are at increased risk to develop diabetes mellitus (DM) compared to their able-bodied counterparts, likely due to body composition alterations and autonomic nervous system dysfunction. These factors are more pronounced in persons with tetraplegia (TP) versus paraplegia (PP), however, the effect of level of injury (LOI) on DM incidence is largely unknown. Therefore, the objective is to examine the effect of LOI on DM incidence in persons with SCI.

**Design:** Retrospective longitudinal cohort study of veterans with SCI.

**Methods:** We obtained electronic record data on age, sex, race/ethnicity, LOI and HbA1c concentration from January 1^st^ 2001 through December 31^st^ 2021. Cox proportional hazard regression analyses were used to assess the association between LOI, DM and all-cause mortality.

**Results:** Among 728 non-diabetic veterans with SCI (350 TP/ 378 PP, 52±15 years, 690 male/38 female) 243 developed DM, of which 116 with TP and 127 with PP. Despite chronological variations between TP and PP, DM risk over the entire follow-up did not differ among the groups (hazard ratio (HR): 1.06, 95% CI: 0.82 - 1.38). Mortality was higher in TP versus PP (HR: 1.40, 95% CI: 1.09 – 1.78). However, developing DM did not increase the risk of death, regardless of LOI (HR: 1.07, 95% CI: 0.83 – 1.37).

**Conclusions:** In this cohort of veterans with SCI, the level of injury had minimal effect on long-term DM development but increased mortality as previously reported.

**Significance statement:** Persons with spinal cord injury are at increased risk for developing diabetes mellitus (DM); however, the effect of level of injury is unclear. In the current study using an electronic health record system we found that the incidence of DM is similar between persons with a high (i.e., tetraplegia) versus low (i.e., paraplegia) lesion level, and that developing DM does not affect the risk for mortality. Additionally, our findings suggest that baseline glycemic level (such as HbA1c), unlike baseline body mass index, is a strong predictor of DM development in this population. Further studies are warranted to explore pathophysiological factors responsible for DM development among patients with higher and lower LOI to develop targeted preventive and therapeutic strategies.

## Introduction

Spinal cord injury (SCI) is a debilitating condition that affects ∼21 million cases worldwide, with a global incidence of ∼1 million per year (1). Recent advances in acute medical care have resulted in a decline in mortality in the early phase post-SCI (2). In an increasingly aging SCI population, the consequences of metabolic disorders, such as diabetes mellitus (DM) and related complications like cardiovascular disease, are now among the most prevalent causes of death (2).

A cross-sectional comparison of 100 veterans with SCI and 50 able-bodied (AB) veteran controls demonstrated that those with SCI have a 3-fold greater risk of abnormal glucose tolerance than AB controls (3). The Canadian Community Health Survey (N: 297 SCI versus 60,381 AB)(4) reported a 150% increase in the risk of T2D after SCI. Adjustment for known T2D risk factors including smoking, hypertension, age, gender, body mass index (BMI), physical activity, alcohol, and dietary intake/composition did not change DM risk in this cohort (4). A greater risk of DM after SCI has also been reported using a nationwide insurance claims database from the USA (N: 9,081 SCI versus 1,474,232 AB)(5) and the Taiwan National Health Insurance Research Database (N: 47,916 SCI versus 191,664 AB)(6). In a national survey of veterans with SCI (N: 3721)(7) the prevalence of DM in those with SCI was 3-fold higher than in those without. Further, veterans with SCI and DM were at higher risk to develop micro- and macrovascular complications compared to veterans having DM without SCI (7).

And yet, the factors through which SCI exacerbates DM development risk is not fully understood. While reduced physical activity levels, muscle atrophy, and increased visceral adiposity after SCI can contribute to glucose abnormalities after SCI (8), less-characterized factors such as autonomic dysfunction-related changes in glucose metabolism and energy homeostasis may also play a role (3, 9). Tetraplegia (TP) causes a greater disruption to the autonomic nervous system as well as more severe muscle atrophy and impaired physical capacity compared with paraplegia (PP)(10). It is therefore plausible that glucose abnormalities are worse in TP than PP. Despite a well-documented increased risk of DM after SCI in general, the role of level of injury (LOI) on glucose abnormalities in this population remains unclear. Previous studies, using oral or intravenous glucose tolerance tests, have shown that persons with TP have impaired glucose tolerance and insulin action compared to those with PP (11, 12). However, these studies were cross-sectional and included a small sample size, underscoring the need for longitudinal investigations with a long follow-up period in a large sample of persons with SCI.

Thus, using a retrospective longitudinal design with a 21-year follow-up, we examined the effect of LOI on DM incidence in a single-center cohort of veterans with SCI. Further, we explored the effect of DM on all-cause mortality as well as the predictive value of baseline candidate risk factors for DM and mortality in this cohort. It was hypothesized that TP was associated with a higher DM incidence compared with PP. Also, we anticipated that DM increased the risk of all-cause mortality in this population independent of LOI.

## Methods

### Study design

We conducted a retrospective longitudinal cohort study of Veterans Affairs (VA) patients with SCI receiving care within the South Texas Veterans Health Care System (STVHCS) between Jan 2001 and Dec 2021. The institutional review board of the University of Texas Health Science Center at San Antonio provided waiver of all aspects of ethics approval. The Veterans Health Administration (VHA) electronic health records (EHR) (VHA corporate data warehouse) were searched for all veterans, who (1) were diagnosed with SCI based on International Classification of Diseases 9^th^ or 10th Revision (ICD-9 or ICD-10) diagnosis codes for PP (ICD-9 = 344.1, or ICD-10 = G82.2X) or TP (ICD-9 = 344.0X, or ICD-10 = G82.5X) in their VHA EHR; and (2) had at least one outpatient visit or inpatient stay in the STVHCS SCI clinic during at least two different calendar years between Jan 2001 and Dec 2021.

Subjects were excluded if their records had no valid diagnostic code for type of SCI or any records of outcome of interest (HbA1c or serum glucose concentration) (Fig. 1). The subjects were then stratified into having PP or TP based on ICD codes as noted above.

**Fig. 1.**
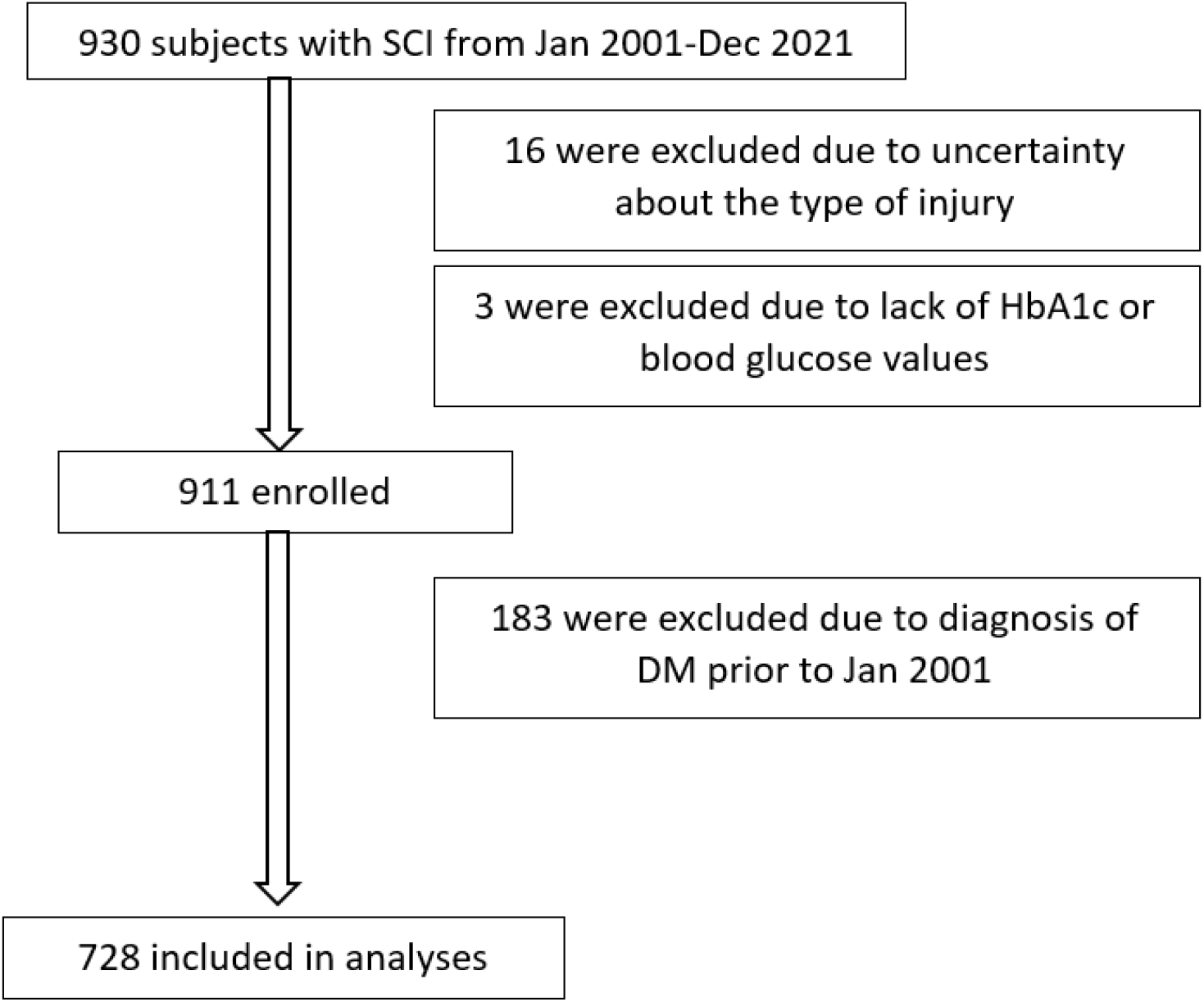
Consort chart of subjects enrolled and included in the analyzed data set.

### Outcome measures

The primary endpoint was the time to new diagnosis of DM based on LOI (i.e., TP vs PP). Secondary endpoints were the effect of LOI and DM on mortality from any cause as well as the relationship between baseline characteristics and the incidence of DM and mortality.

Using standardized data definitions and programming specifications a dataset was obtained for each eligible subject within STVHCS using the VHA corporate data warehouse. Data included baseline demographic characteristics (sex, age, and ethnicity), BMI, laboratory data related to glycemic control (HbA1c and serum glucose concentration), pharmacy data related to anti-diabetic medications, outpatient diagnosis, and survival status.

Diabetes mellitus was defined as one or a combination of the following conditions: (i) HbA1c measure ≥ 6.5%; (ii) random plasma glucose concentration ≥ 200 mg/dl; (iii) taking anti-diabetic medication (e.g.: metformin, insulin, amylinomimetics, DPP-4 inhibitors, GLP-1 receptor agonists, meglitinides, SGLT-2 inhibitors, sulfonylureas, or thiazolidinediones). Persons with SCI in our cohort were typically seen every 6 -12 months. DM screening occurred annually in all patients with blood draws. While random glucose was obtained, HbA1c was used as diagnostic since only one blood sample was obtained from patients and there was heterogeneity in fasting status. Further, the problem list was not always generated by the health care practitioner, therefore, presence or absence of DM in the problem list was not a reliable source for DM diagnosis in our search algorithm. If DM occurred prior to the first visit in the SCI clinic (the “baseline” date), or up to 60 days after the first SCI visit, the patient was classified as “DM present at baseline” and excluded from the regression analyses. The glycemic indices (i.e., HbA1c and random glucose concentration) were collected from the EHR for the first SCI visit and at the time of DM diagnosis (when indicated), based on the specimen collected closest in time, within 60 days of each timepoint. Time until DM development or death was calculated from the first encounter in each SCI subjects until the date of each outcome or end of the observation period. The maximum possible observation time was 21 years (January 1, 2001 – December 31, 2021)

### Statistical analysis

Baseline characteristics were compared based on LOI using ANOVA or chi-squared tests when indicated. The incidence of DM and all-cause mortality between TP and PP were calculated using Kaplan-Meier estimator over 21-year follow-up period. The DM incidence and all-cause mortality were then compared between subjects with TP versus PP using a Cox proportional hazard model for DM and death, which were adjusted for several baseline covariates (age, sex, and race/ethnicity). The effect of baseline characteristics on DM incidence or mortality was also calculated with and without adjusting for LOI using the likelihood ratio test. Subjects were censored for lost to follow-up or death. All analyses were performed by an independent VA statistician (MM), using SAS version 9.4 and statistical significance was accepted at *p*<0.05 or when the 95% confidence interval did not include 1.

## Results

### Subjects

A total of 930 subjects with SCI were identified at baseline; 19 were excluded due to an uncertainty about the type of injury or lack of baseline glycemic indices (Fig 1). Out of the 911 subjects at baseline, 471 had PP and 440 TP (Table 1). More than 90% of the subjects were male, reflecting the veteran population(13). Subjects with PP were slightly younger (*p* <0.05) and had a higher BMI (*p*<0.001) than those with TP at baseline. The baseline HbA1C and random glucose concentration were similar among the two groups, and so was the prevalence of DM (TP: 20.5%, PP: 19.7%). The majority of the cohort was White, followed by Hispanic, then Black. Compared to PP, there was a larger number of Black and smaller number of White persons in TP (*p* <0.05). Subjects without DM at baseline (N = 728) were followed up for a median time of 94 (TP) versus 114 (PP) months (*p* = 0.06).

**Table 1.** Subject characteristics at baseline.

| Baseline Characteristics | Paraplegia (N=471) | Tetraplegia (N=440) |
| --- | --- | --- |
| Age (yrs) | 53.5±15.4 | 55.5±14.4* |
| Gender (N males (%)) | 442 (93.8%) | 421 (95.7%) |
| Race/Ethnicity (N (%)) |  |  |
| Hispanic | 92 (19.5%) | 87 (19.8%) |
| White | 260 (55.2%) | 205 (46.6%)* |
| Black | 50 (10.6%) | 71 (16.1%)* |
| Other | 12 (2.5%) | 11 (2.5%) |
| Unknown | 57 (12.1%) | 66 (15.0%) |
| Diabetes Mellitus (N (%)) | 93 (19.7%) | 90 (20.5%) |
| BMI (kg/m<sup>2</sup>) | 27.7±6.4 | 26.0±5.5** |
| HbA1C (%) | 5.6±1.1 | 5.5±1.1 |
| HbA1C (mmol/mol) | 34.1±7.6 | 33.4±6.4 |
| Glucose # (mg/dl) | 112.2±40.3 | 109.5±33.8 |

Data are presented as mean ± SD unless specified otherwise; # Random plasma glucose concentration; HbA1c: glycated hemoglobin;
\* $p < 0.05$ compared to paraplegia; \*\* $p < 0.001$ compared to paraplegia

### Incidence of diabetes mellitus in subjects with paraplegia and tetraplegia

During the 21-year follow-up, 243 subjects developed DM, of which 116 with TP and 127 with PP. The median time to a new diagnosis of DM was 39 versus 40 months in TP and PP, respectively (*p* = 0.065). We found a chronological variation in DM incidence based on the level of injury (Fig. 2): (1) In the first five years, there was no difference between TP and PP; (2) From 5 to 10 years, the incidence of DM increased to greater extent in TP compared to PP with the largest difference observed in year 7 (hazard ratio (HR): 1.33, 95% CI: 0.99 – 1.79); (3) After 10 years, the incidence in both groups converged towards the end of the follow-up period. The risk of DM calculated for the entire follow-up period did not differ between the two groups (3.64 per 100 subject-years in TP vs. 3.36 per 100 subject-years in PP). Adjusting for baseline age, sex and race/ethnicity did not change the lack of association between LOI and DM for the 21-year follow-up period (HR: 1.08, 95% CI: 0.84 – 1.39; adjusted HR: 1.06, 95% CI: 0.82 - 1.38).

**Fig 2.**
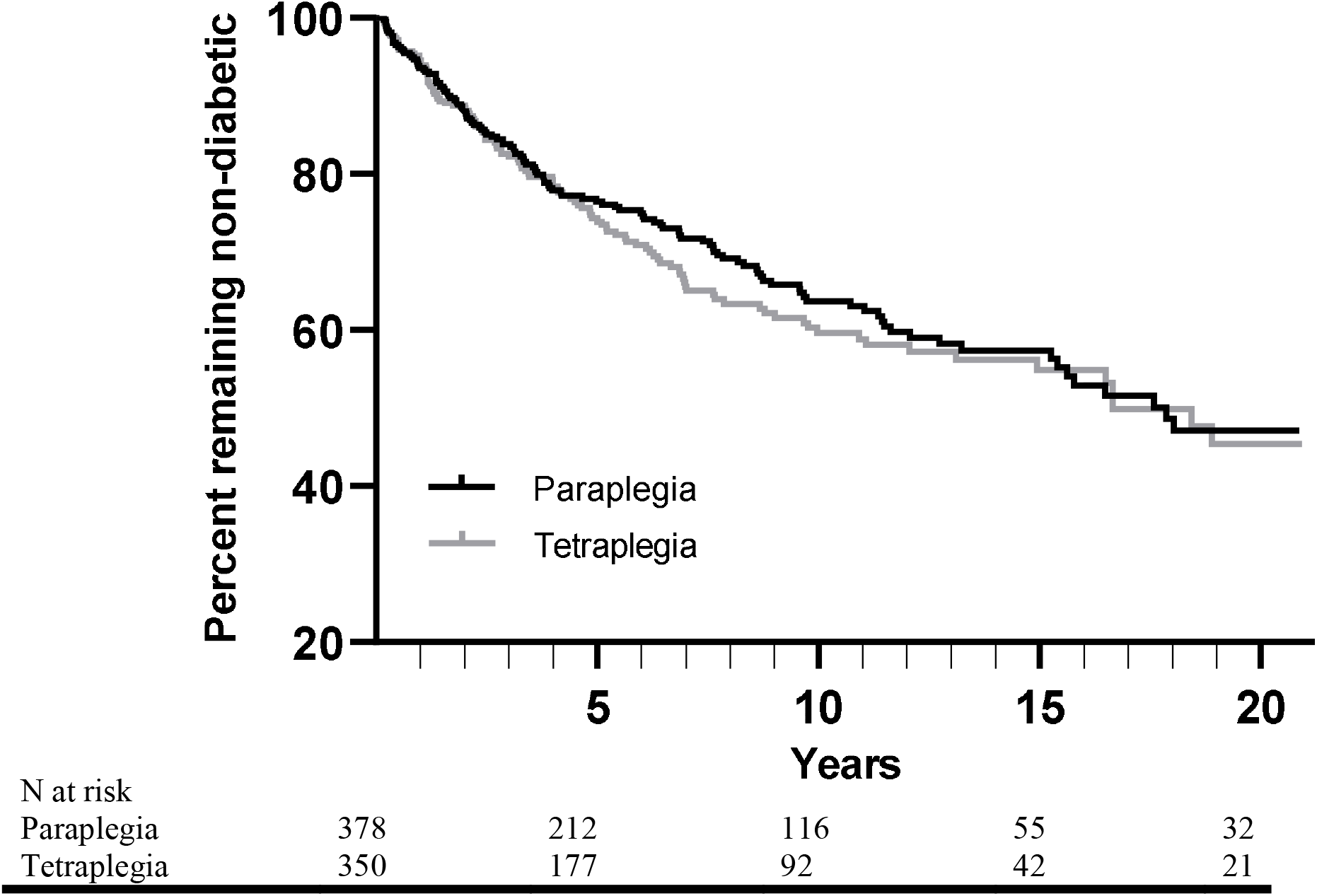
Rates of diabetes mellitus in subjects with tetraplegia and paraplegia during the 21-year follow-up.

**Fig. 3.**
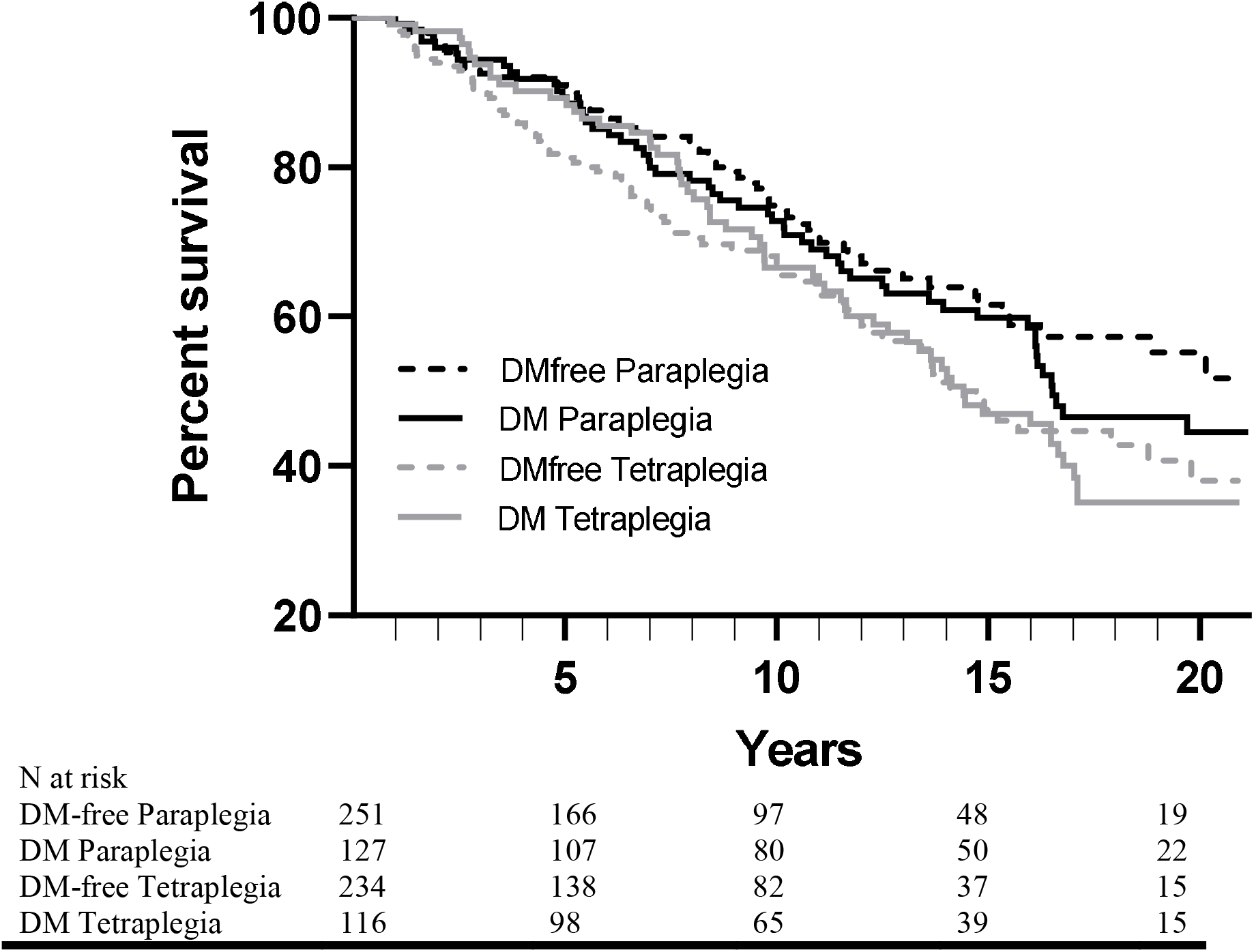
Rates of mortality in subjects with and without diabetes mellitus developed during the 21-year follow-up. DM: diabetes mellitus

### Mortality rates in subjects with paraplegia and tetraplegia

A total of 260 SCI subjects died during the 21-year follow-up, of which 140 with TP and 120 with PP. There was a trend of shorter time to death in TP than PP (median time of 92 versus 99 months; *p* = 0.060). Persons with TP had a ∼40% increased risk of mortality compared to persons with PP (HR: 1.40, 95% CI: 1.09 – 1.78). However, the effect of LOI on mortality was reduced after adjusting for baseline age, sex and race/ethnicity (adjusted HR: 1.19, 95% CI: 0.92 - 1.53). In contrast, adjusting for DM status, those with and without new DM diagnosis during the follow-up period, had no additional influence on the risk of mortality (HR: 1.07, 95% CI: 0.83 – 1.37).

### Baseline characteristics that predict diabetes mellitus development and mortality in subjects with spinal cord injury

Older age, HbA1C, and BMI at baseline increased the risk for DM development (*p*<0.001, *p*<0.001, *p*<0.05, respectively); although the strongest association with DM risk was found for HbA1c and the weakest for BMI. Females tended to have a lower risk for DM development and those of Black race compared to White race tended to have a higher risk, but neither estimate reached statistical significance (Table 2). Adjusting the estimates by LOI had no additional effect on the outcomes.

**Table 2.** Baseline risk factors for converting to diabetes mellitus and mortality.

|  | **DM**<br>Unadjusted HR (95% CI) | **Mortality**<br>Unadjusted HR (95% CI) |
| --- | --- | --- |
| Age (baseline) | 1.028 (1.019-1.037)** | 1.070 (1.059-1.081)** |
| Sex (F vs M) | 0.549 (0.281-1.074) | 0.477 (0.291- 0.782)<sup>#</sup> |
| BMI (baseline) | 1.026 (1.005 – 1.047)<sup>#</sup> | 0.960 (0.936 – 0.984)* |
| HbA1c (baseline) | 1.779 (1.572 – 2.012)** | 1.087 (0.879 – 1.344) |
| Glucose (baseline) | 1.023 (1.017 – 1.028) <sup>#</sup> | 1.009 (1.002 – 1.016) <sup>#</sup> _ |
| Race/Ethnicity |  |  |
| Black vs. White | 1.392 (0.960-2.018) | 1.116 (0.701-1.776) |
| Hispanic vs. White | 0.921 (0.656-1.293) | 0.875 (0.591-1.296) |

Abbreviations. DM: diabetes mellitus, HR: hazard ratio, LOI: level of injury, HbA1c: glycated hemoglobin, F: female, M: male, TP: tetraplegia, PP: paraplegia, Glucose: plasma concentration of random glucose. <sup>#</sup> $p<0.05$ , \* $p<0.01$ , \*\* $p<0.0001$

As expected, older age at baseline increased the risk of mortality (*p*<0.001), while a higher BMI at baseline was associated with a reduced mortality risk (*p*<0.01). Finally, females had a reduced risk of mortality compared with males (*p<*0.05) (Table 2). None of the mortality predictors were affected by adjustment for LOI (*p*>0.05).

## Discussion

Cross-sectional data indicate that adverse cardiometabolic effects of SCI, including DM risk factors, are exaggerated in those with a higher LOI (9, 12). Here we investigated the effect of LOI on the incidence of DM in a retrospective, longitudinal single-center veteran cohort study. The risk of DM development estimated over the 21-year follow-up period did not differ among those with TP versus PP. However, there was a chronological difference in DM development based on LOI as subjects with TP had a ∼30% higher risk compared to those with PP in Year 7, which dissipated thereafter. Also, there was a trend of shorter time to new DM among TP versus PP. Altogether, these findings suggest that the diabetogenic effect of LOI is complex.

Consistent with previous reports, in our cohort, subjects with TP had a higher mortality than PP, but the risk of mortality was not changed by DM development. We further found that subjects with a higher HbA1c concentration at baseline were more likely to develop DM but mortality was not affected by variations of this parameter. The baseline BMI had a small but statistically significant positive association with DM risk as well as reducing mortality among subjects with SCI.

In our cohort, ∼20% of the individuals with SCI had DM at baseline, detected by random glucose and HbA1C. While we did not include AB controls in this study, baseline prevalence of DM is considerably higher than the ∼10% previously reported in the general USA population (14). Similarly, we found a higher incidence of DM at 4.7 per 100 person-years in our SCI cohort compared to the 0.7-1.3 per 100 person-years that were previously reported in AB middle-aged and older adults (15, 16). Our findings of higher prevalence and incidence of DM in persons with SCI compared with the general population are in corroboration with previous studies (4, 6, 17).

However, in contrast to our hypothesis and findings from previous cross-sectional studies (9, 11, 12, 18), the current longitudinal study does not provide support for a major difference in DM incidence between persons with TP and PP, despite a trend for higher DM risk in TP in the first decade of follow-up as well as a trend of shorter time to new DM in this population.

The previously reported distinct higher risk of impaired glucose tolerance among individuals with TP as opposed to PP (9, 11, 12, 19) has been attributed to the severity of autonomic nervous system disruption, lean/fat mass redistribution (8, 11), inflammation (18) and lipid abnormalities (9) in SCI patients with higher LOI. The loss to follow-up beyond 10 years may have biased the sample in the second part of our observation. Further, a 40% higher risk of mortality in TP than PP and lack of information about the cause of death could have distorted the balance between the two groups at-risk in the latter phase of observational period. A previous study using the same method of retrospective data analysis in persons with SCI (mean age of ∼52 years, ∼64% men) compared to matched AB subjects from the background Taiwanese population, found a ∼30% increased risk of DM in SCI compared to AB (6). The DM incidence in the SCI cohort was higher in males than females, and those with a higher age. Importantly, however, individuals with a thoracic or lumbar lesion (PP) had a *higher* risk of DM compared to those with a cervical lesion (TP); a finding that the authors related to a lower life expectancy in individuals with TP than PP (6), which has been well documented (2).

While data on the change in BMI over the follow-up period is missing in the current study, previous reports indicated that subjects with PP had a larger increase in BMI over the follow-up period than those with TP (20, 21). In our study, baseline BMI was only a weak predictor of DM. This observation is likely due to underestimation of obesity in SCI population by using conventional measures of adiposity, such as BMI. However, a within-person BMI increase that is typical of persons with PP versus TP(22), likely reflects increased obesity over time, which can adversely affect glucose metabolism, insulin secretion and insulin action. Thus, a potential increased adiposity in PP may have overridden the protective effect of a lower LOI in our cohort (23). Indeed, cross-sectional studies showing a deteriorated glycemic profile in TP compared with PP have often matched total fat percentage between both groups (9, 18). We infer from the conflicting data provided by previous cross-sectional and our current longitudinal studies that the effect of LOI on DM risk is likely dependent on factors yet unexplored, such as time since injury and adiposity.

As noted earlier, the present study underscores the previously reported limited value of BMI as a predictor for metabolic disease in persons with SCI (24, 25). On the other hand, Rajan et al. (19) showed that a diagnosis of overweight or obesity based on BMI is associated with increased DM risk among persons with SCI. Although, in that study, a single unit increase in BMI at baseline was associated with only a 4% DM risk increase (19); similar to the 3% observed in the current study. In comparison, while a 10 unit increase in BMI (e.g., 20 kg/m2 to 30 kg/m2) raises the risk by ∼30%, a one unit increase in HbA1c (e.g., 4.5% to 5.5%) at baseline in the current cohort increases the risk of DM development by nearly 80% (Table 2). Future studies may therefore benefit from including dual energy X-ray absorptiometry (DEXA) or a more accessible outcome such as waist circumference to provide more insight into the role of body composition in glucose abnormalities in persons with SCI (24, 25, 26).

In corroboration with previous studies (2, 27), all-cause mortality in our cohort was higher than that reported in the general population and was higher in TP versus PP (27, 28). However, in contrast to our hypothesis and observations in the general population (29, 30), mortality rates were not different between those who developed glucose abnormalities and those who remained DM-free during the follow-up period. Although the proportion of metabolic disorders as a cause of death has risen in the past decades, respiratory complications are still the leading cause in persons with SCI, followed by infectious disease, cancer and heart disease (2). One of the limitations of our analysis is that the data from specific cause of death in our cohort is lacking. Thus, it remains to be further investigated whether DM associated complications impact mortality in relation to other SCI-related conditions (respiratory and vascular complications, as well as infections) that are likely to be higher in TP than PP (10).

### Practical implications

Glucose intolerance and insulin resistance is increasingly recognized as the pillar on which many chronic diseases rest (36), linking DM to the development of cardiovascular disease, several types of cancer and Alzheimer’s disease (37). Although DM was not associated with increased mortality risk in our studied cohort, the high incidence of DM found here is sufficient to call for further studies to investigate pathophysiological differences among persons with TP and PP, which in turn can help with the development of personalized preventive and treatment strategies that are specific to the lesion level. The similar DM incidence in TP and PP found in the current study suggests that resources for such endeavors should be divided equally among these SCI sub-populations.

### Limitations

Several limitations of the present study need mentioning. First, DM diagnosis was based on one rather than two blood sample collections (35) given the nature of retrospective study and not having access to more than one measurement per 6-12 months. This may have resulted in an overestimation of the DM incidence; but this effect is unlikely to have been different between TP and PP. Furthermore, we were unable to obtain data on lesion completeness. Asymmetry in this factor between both groups may partly explain the lack of a difference in DM incidence between TP and PP (6, 9). For instance, in aging Western societies, the SCI demographic has shifted from predominantly young individuals with complete PP to older persons with incomplete TP (41).

Case in point, a separate analysis on the patients that visited our center within the last year (n=384), indicated that ∼75% of patients with TP (n=141 of 182) vs. ∼50% of patients with PP (n=103 of 203) have an incomplete lesion. Also, we could not obtain data on the time since the injury. However, given the veteran health care system with a standardized trajectory from trauma hospital to inpatient and subsequently outpatient care, it is likely that the time interval between the time of injury and first encounter was similar between PP and TP and no more than one year from injury. Additionally, as the DM diagnosis was based on HbA1c and random glucose concentration only, were unable to differentiate between type 1 and type 2 DM. However, considering the relatively old age of the studied cohort, it is prudent to suggest that the proportion of type 1 DM was small. Finally, the studied cohort of American veterans consisted of predominantly males and has its own unique characteristics. As such, the findings may not be directly transferable to female cohorts or those with a different professional background.

## Conclusions

Although there was a chronological trend of higher initial DM development in persons with TP than those with PP, overall incidence of DM did not differ between both groups during a 21-year observational period. Mortality was higher in TP compared with PP, but developing DM was not associated with a higher risk of death in this SCI cohort. Finally, the present longitudinal study provides support for the continued use of HbA1c to identify people at risk for DM, while the HbA1c threshold for DM-risk or cardiometabolic complications in this population remains to be investigated.

## Data Availability

All data produced in the present work are contained in the manuscript.

## Acknowledgements

This material is the result of work supported with resources and the use of facilities at the Audie L. Murphy Veteran’s Affairs Hospital in San Antonio, TX.

## Data availability

All data produced in the present work are contained in the manuscript.

